# Personalized Prediction of Disease Activity in Patients with Rheumatoid Arthritis Using an Adaptive Deep Neural Network

**DOI:** 10.1101/2020.09.03.20168609

**Authors:** Maria Hügle, Ulrich A. Walker, Axel Finckh, Rüdiger Müller, Gabriel Kalweit, Almut Scherer, Joschka Boedecker, Thomas Hügle

## Abstract

**Background:** Rheumatoid arthritis (RA) lacks reliable biomarkers that predict disease evolution on an individual basis, potentially leading to over- and undertreatment. Deep neural networks learn from former experiences on a large scale and can be used to predict future events as a potential tool for personalized clinical assistance.

**Objective:** To investigate deep learning for the prediction of individual disease activity in RA.

**Methods:** Demographic and disease characteristics from over 9500 patients and 65.000 visits from the Swiss Quality Management (SCQM) database were used to train and evaluate an adaptive recurrent neural network (AdaptiveNet). Patient and disease characteristics along with clinical and patient reported outcomes, laboratory values and medication were used as input features. DAS28-BSR was used to predict active disease and future numeric individual disease activity by classification and regression, respectively.

**Results:** AdaptiveNet predicted active disease defined as DAS28-BSR>2.6 at the next visit with an overall accuracy of 75.6% and a sensitivity and specificity of 84.2% and 61.5%, respectively. Regression allowed forecasting individual DAS28-BSR values with a mean squared error of 0.9, corresponding to a variation between predicted and true values at next visit of 8%. Apart from DAS28-BSR, the most influential characteristics to predict disease activity were joint pain, disease duration, age and duration of treatment. Longer disease duration, age >50 years or antibody positivity marginally improved prediction performance.

**Conclusion:** Deep neural networks have the capacity to predict individual numeric disease activity in RA. Low specificity remains challenging and might benefit from alternative input data or outcome targets.

## INTRODUCTION

Rheumatoid arthritis (RA) is a chronic inflammatory disorder in which disease activity fluctuates over time. The advent of targeted synthetic and biologic medication, along with early and treat-to-target strategies have substantially improved patient care. However, sustained remission still is only achieved in around 30% indicating room for improvement either by new drugs or alternative treatment strategies^1^. EULAR/ACR recommendations suggest treatment modification after three to six months if the set target is not reached, regardless of the presence or absence of individual risk factors for poor outcome^2^. Given the increasing number of available drug combinations, the delay in finding the best individual treatment can be substantial. Despite the advent of new biomarkers, their practical role to predict individual chances of good therapeutic response remains limited^3,4^. There are also no clear recommendations on treatment de-escalation in case of stable disease^5^ despite disease activity-guided dose optimisation of biologic being efficient and cost-effective^6^. In other words, over- or undertreatment in RA is common, potentially resulting either in destructive disease flares or unnecessary side effects and costs^7^.

Machine learning (ML) is a relatively new approach for disease detection, disease stratification and disease prediction both in at risk populations and established disease^8^. Using data from electronic medical records (EMR), ML has successfully predicted RA flares in a small number of RA patients by a random forest, as basic machine learning method^9^. Norgeot et al. applied deep learning^10^ (DL) as a newer subfield of ML to EMR data in 820 RA patients for the prediction of disease activity by classification^11^. To predict the category of low disease activity, a remarkable AUC score of 0.91 was achieved in a test set of 116 patients. Medication in this setting could not be assessed due to the incomplete EMR dataset. Using the Swiss Quality Management (SCQM) database^12^ for rheumatic diseases, we recently described a novel adaptive deep neural network being superior to conventional DL architectures in the prediction of disease activity in a larger dataset of 9500 RA patients^13^. The study presented here aims to characterize this deep neural network in RA patients, to forecast individual disease activity both categorically and numerically as a potential tool for assistance in clinical decision.

**Table 1:** Performance of an AdaptiveNet model in RA patients for prediction of active disease in training sets from different patient subsets. Accuracy, sensitivity, specificity, and area under the curve (AUC) indicate performance for classification, mean squared error (MSE) for regression.

|  | Accuracy | Sensitivity | Specificity | AUC | MSE |
| --- | --- | --- | --- | --- | --- |
| All patients (Max. observation time 5 years) | 0.7558 | 0.8417 | 0.6149 | 0.7283 | 0.905 |
| Age $\geq 50$ years | 0.76 | 0.8513 | 0.5924 | 0.7219 | 0.879 |
| Age < 50 years | 0.742 | 0.8035 | 0.6679 | 0.7357 | 0.989 |
| Disease duration < 3 years | 0.7077 | 0.8348 | 0.5658 | 0.7003 | 1.058 |
| Disease duration $\geq 3$ years | 0.7599 | 0.8393 | 0.6232 | 0.7312 | 0.887 |
| Rheumatoid factor positive | 0.7691 | 0.8581 | 0.6056 | 0.7319 | 0.916 |
| Rheumatoid factor negative | 0.7172 | 0.7773 | 0.641 | 0.7092 | 0.871 |
| Anti-CCP negative | 0.7461 | 0.8227 | 0.6262 | 0.7245 | 0.922 |
| Anti-CCP positive | 0.7415 | 0.784 | 0.6793 | 0.7316 | 0.851 |
| Male | 0.7578 | 0.7796 | 0.7213 | 0.7504 | 0.975 |
| Female | 0.7548 | 0.8578 | 0.5645 | 0.7111 | 0.884 |

## METHODS

### Dataset

The dataset used is the Swiss Clinical Quality Management in Rheumatic Disease (SCQM) registry, a national multicenter database containing longitudinal data from clinically diagnosed RA patients. Patients are followed-up with one to four visits yearly including clinical, radiographic and patient reported outcome data. Characteristics of the database are described elsewhere^12^. The collection of patient data for the SCQM register was approved by a national review board and all individuals willing to participate sign an informed consent form before enrolment, in accordance with the Declaration of Helsinki.

### Input features and prediction target

To predict disease activity, we used DAS28-BSR as target variable and considered only visits with complete scores. We used age, gender, weight, disease duration, BSR, CRP, swollen joint count, painful joint count, rheumatoid factor, anti-CCP, treatment, smoking status, HAQ, morning stiffness, EuroQol, disease activity and pain level as potential predictors. For antirheumatic therapy, we used the individual drugs, as well as broader drug categories (biologic, csDMARDs and prednisone dose strata) and duration of therapy since adjustment. For training and evaluation of the predicted target variable we considered follow-up visits between 1 month and 1 year. As input features, we considered the visit and medication data of the last 5 years.

### Classification and regression

For classification, we defined two disease states, active disease (DAS28-BSR > 2.6) and remission (DAS28-BSR ≤ 2.6) at next visit. Prediction performance was measured by accuracy, sensitivity, specificity and area under the curve (AUC) score. In order to predict numeric values of the target variable (DAS28-BSR), we applied a regression model and predicted the expected change of DAS28-BSR to the subsequent visit. Performance was measured by mean squared error (MSE) as an estimator of the deviation between the estimated and actual values. To evaluate the models, we split the dataset into a training set and a validation set by using 5-fold cross-validation.

### Modelling

Classification and regression was performed with AdaptiveNet, a dynamic and recurrent deep neural network architecture, designed for chronological clinical data^13^. In short, AdaptiveNet encodes all former clinical events of a patient (here: visits and medication adjustments) to the same latent space using multiple fully-connected encoder networks in order to align the corresponding output vectors (Figure 1). Sorted lists of these encoded clinical events are pooled by a long short-term memory^14^ (LSTM) to compute a fixed-length encoding, representing the 5-year patient history. The final output is computed by a fully-connected network module, using the encoded patient history and additional features containing general time-independent patient information as input. Preprocessing, architecture, learning rate, optimizer and batch size are described in Hügle et al.^13^. We used loss of MSE for regression and binary cross-entropy for classification. To estimate feature importances, we additionally trained a random forest^15^ with a maximum depth of 10.

**Figure 1:**
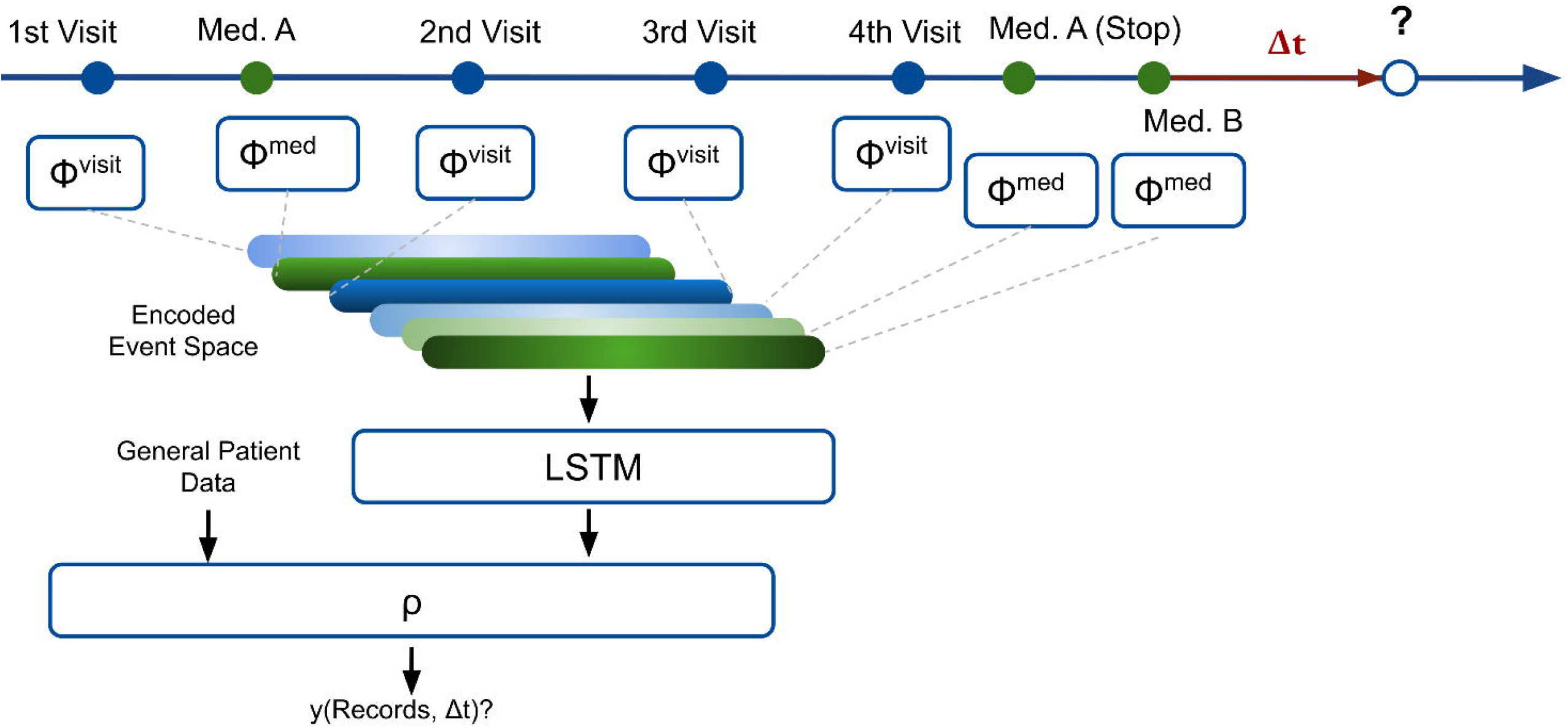
Deep neural network architecture (AdaptiveNet). All visits and medication adjustments are projected to latent vectors of the same size using encoder networks □_visits_ and □_meds_. Latent vectors are sorted according dates and fed into a long short-term memory (LSTM) to create a latent vector describing the full patient history. The final prediction is computed by the network module ρ, exploiting the patient history with general patient information.

## RESULTS

### Categorical prediction of disease activity by classification

In total, 28.601 visits with corresponding disease activities were extracted. Over a maximal observed history length of 5 years, patients had 6.3 (±5.3) visits and 2.5 (±2.7) medication adjustments. For the classification task DAS28-BSR>2.6 at next visit, in mean taking place 8.1 +-2.9 months after the visit of interest, AdaptiveNet had an accuracy of 75.6%, a sensitivity of 84.2% and a specificity of 61.5% (Table 1). The Receiver Operating Characteristic Curve (ROC) is shown for all patients (Figure 2a) and different patient subgroups (Figure 2b-f). The DL performance was higher in patients with longer disease duration (Figure 2b), male gender (Figure 2c) and positive rheumatoid factor (Figure 2e). Data from patients aged >50 years achieved a higher accuracy and sensitivity to predict active disease than aged <50. Anti-CCP positive status only showed a marginally better learning performance (AUC 0.73 vs. 0.72) for this task (Table 1).

**Figure 2:**
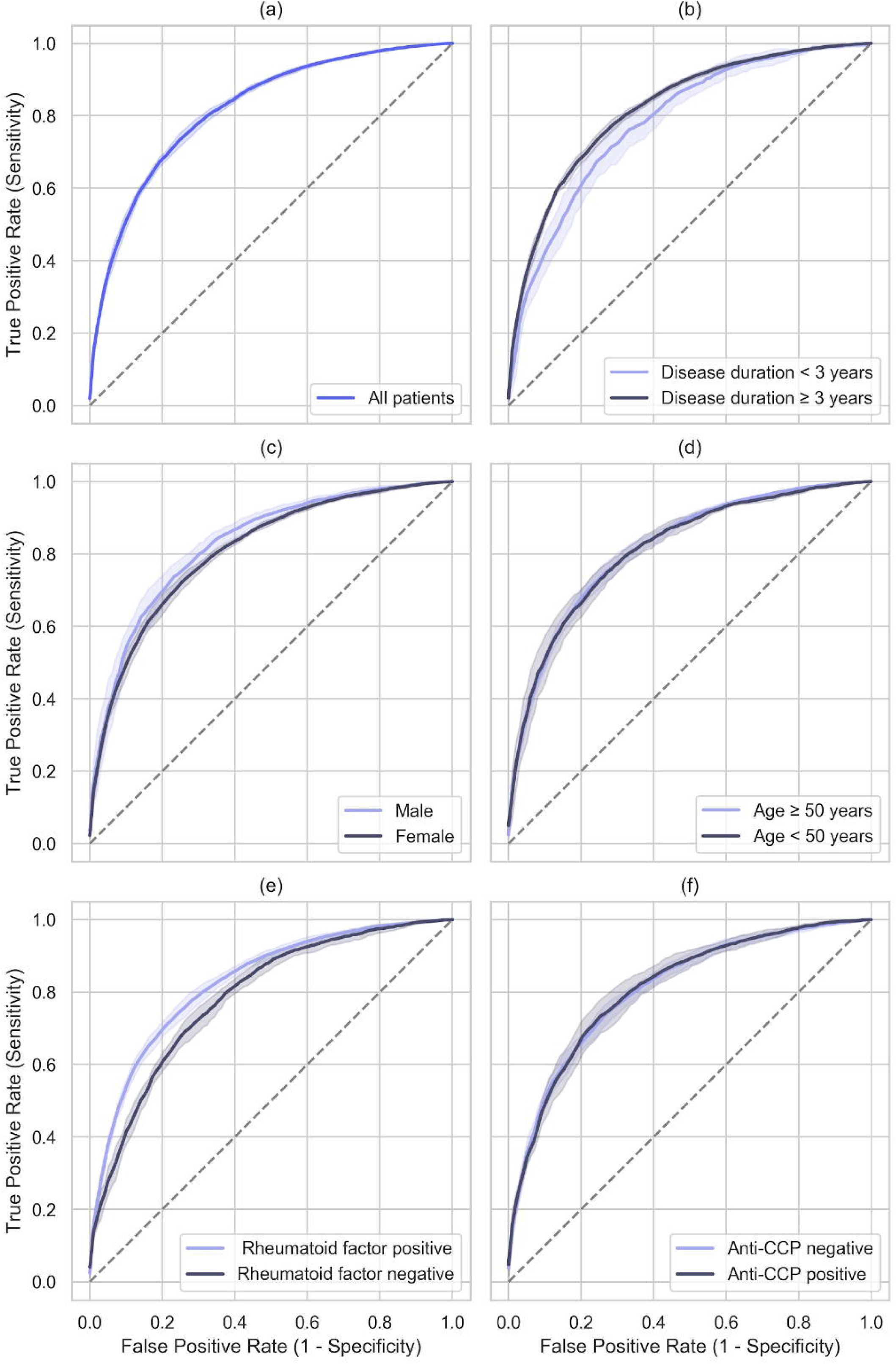
Classification performance of AdaptiveNet to predict active disease (DAS28- BSR>2.6) in different patient subsets shown by Receiver Operating Characteristic Curves. Accuracy and corresponding AUCs are indicated in table 1.

### Numerical prediction of disease activity by regression

AdaptiveNet was applied to predict the numerical DAS28-BSR value at the next visit by regression on an individual level. When trained on data from all patients, we obtained an overall MSE of 0.9 which corresponds to a 8% deviation between estimated and real DAS28-BSR values. Figure 3 shows two patient examples with individual forecasts of DAS28-BSR values over time. A general capacity of the model to predict disease flares as well as response to treatment could be demonstrated. Predicted DAS28-BSR amplitudes during flares were lower than real values and smaller variations of disease activity were not predictable. We obtained better results for patients with disease duration >3 years, age >50 and positive anti-CCP antibodies (Table 1). In contrast to classification, regression had lower MSE values and thus performed better in female and RF-negative patients.

**Figure 3:**
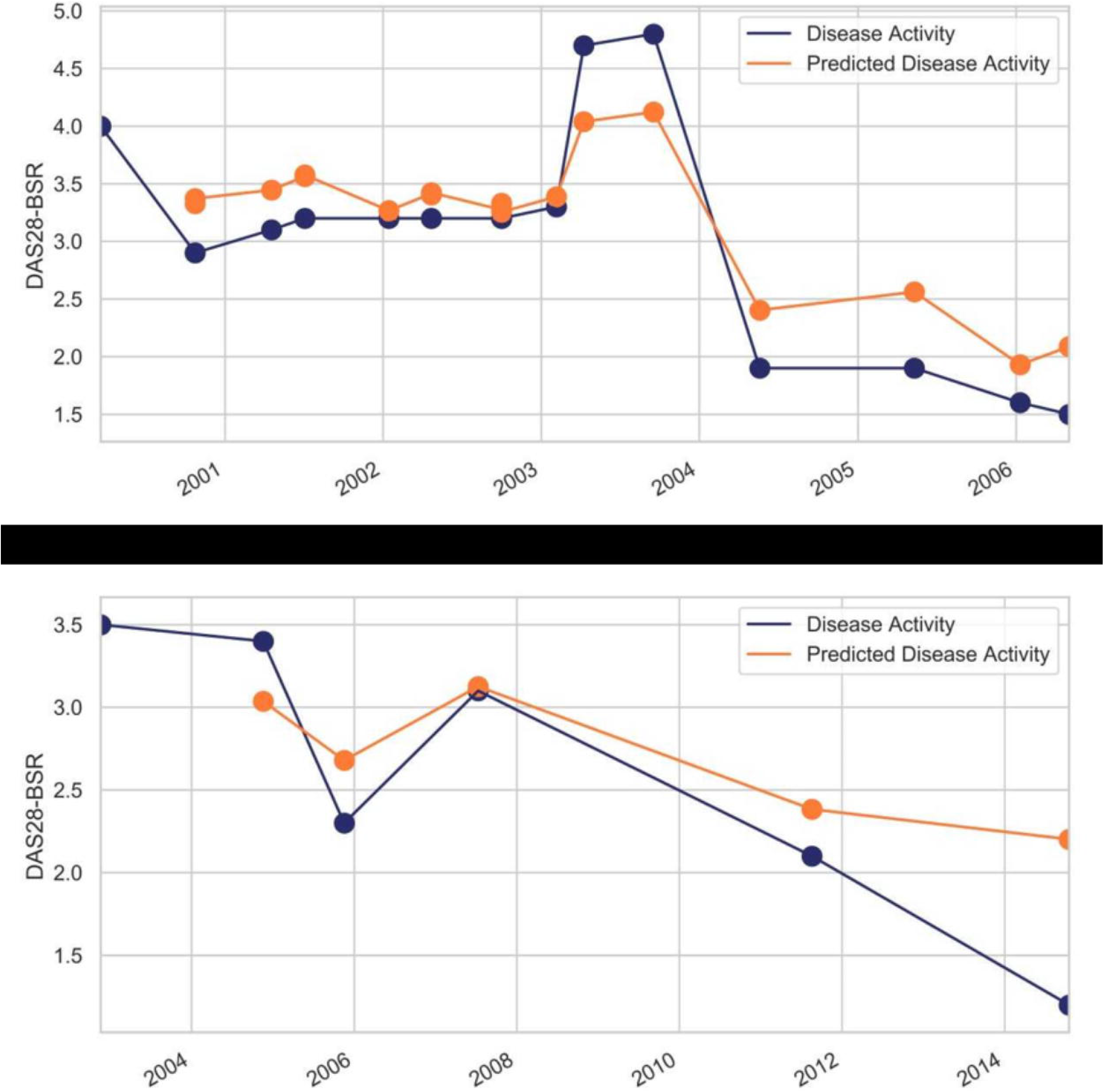
Examples of true disease activity and corresponding predictions of AdaptiveNet by regression analysis. Predictions are made step to step from the current to next visit.

### Feature importance

Feature importance was determined to define the relative importance of variables for disease prediction (Supplement 1). Apart from the target variable itself, the number of painful joints, longer disease duration and age turned out to be the most relevant factors, followed by medication in general, time point of last medication adjustment, number of swollen joints, and HAQ. The importance of medication type (csDMARD vs. bDMARD or corticosteroids) for the prediction of DAS28-BSR was only marginal. Infliximab, tozilizumab and steroids had a slightly higher influence than csDMARDs or other bDMARDs in predicting disease activity.

## DISCUSSION

This study demonstrates a comprehensive classification and regression analysis using deep learning on a large RA dataset. This ML technique allowed to generate individual predictions of subsequent DAS28-BSR values, as shown in Figure 3, instead of disease states alone, which might facilitate the application of DL predictions in the clinic. Thus, DL could foster personalized medicine, e.g. to assist in setting control intervals, and for (de)escalation of treatment. As a further new finding we describe that long disease duration, age>50 and antibody positivity increase the predictability of the active disease. This information could be of importance e.g. for patient selection in future ML-assisted clinical trials. In contrast to classification, the prediction of numeric DAS28-BSR by regression performed better in female than male and anti-CCP positive than in anti-CCP negative patients. We postulate that this difference is due to the fact that classification tasks are prone to overfit to the old class, i.e. predicting no change to the previous situation. In this case, this means that patients in remission for a long period likely will stay in remission, or vice versa, patients resistant to multi-line treatment will more likely remain in active disease. This might also apply to similar classification results shown in other studies^11^. We also performed the classification task to predict DAS28-BSR in- or decrease at next visit (data not shown). This task had a lower accuracy, likely because small fluctuations of DAS28-BSR values are more difficult to predict. Therefore we speculate that that regression could be a more adequate prediction tool for ML-assisted care than classification. Variations of 8% between estimated and real DAS28-BSR values seem acceptable results to implicate disease prediction in the clinical practice. Independently, variable and noisy data in medical databases remain a major challenge, both in EMR and in registry datasets. Advanced architectures like AdaptiveNet improve the performance of prediction in such data compared to classical ML methods^13^ e.g. by taking into account the timing between visits and therapy initiation. The relatively low specificity, however, shows further room for improvement. Taking into account larger datasets through -omics or digital biomarker e.g. by wearables and more patient reported outcomes might further improve the results of disease prediction^16^. To some extent surprising, medication was less important for the prediction of disease activity than age or disease duration. The reason for this might be explained by limited effectiveness after multi-line treatments or vulnerability of DAS28-BSR as target variable to confounding factors as e.g. fibromyalgia. The slightly higher performance of infliximab to forecast disease activity is reasonable from a clinical perspective by intravenous application and dose. Whether DL is able to predict drug survival or individual treatment responses needs to be clarified. Further studies also need to investigate the performance of DL using alternative input and target features including other markers for disease activity than DAS28-BSR. Taken together, we are convinced that DL will play an increasing role to improve patient care and to foster personalized treatment and shared-decision making in patients with RA. Prospective trials will be necessary to prove efficacy, safety and cost effectiveness of ML-assisted care in arthritis.

## Data Availability

Data can be obtained by the corresponding author.

---

Supplementary figure 1: Feature importance. The relative importance of variables for prediction of active disease is computed by a random forest, considering features of the last visit and last medication. Drug classes and individual drugs are indicated separately in the lower part.

## ACKNOWLEDGEMENT

A list of rheumatology offices and hospitals that are contributing to the SCQM registries can be found on www.scqm.ch/institutions.

## FUNDING DISCLOSURE

The SCQM is financially supported by pharmaceutical industries and donors. A list of financial supporters can be found on www.scqm.ch/sponsors.

## COMPETING INTERESTS

None.

## CONTRIBUTORSHIP

MH: Study design, deep learning architecture design and analysis. Manuscript preparation. GK: deep learning architecture design and analysis. UAW: collection of data and data analysis and manuscript preparation. RM: collection of data and data analysis. AF: collection of data, data analysis and manuscript preparation. AS: Study design, and analysis. Manuscript preparation JB: deep learning architecture design and analysis. TH: Study design, collection of data, data analysis and manuscript preparation.

## ETHICAL APPROVAL INFORMATION

Ethical approval was obtained the by the local ethical committee. Informed consent was signed by all patients.

## DATA SHARING STATEMENT

Results are summarized in table 1. The deep learning architecture is published in reference 13. Primary dataset is not available online but can be requested via www.scqm.ch and used for projects in line with rules for research and collaboration of SCQM.

## PATIENT AND PUBLIC INVOLVEMENT

Patients will be informed via different channels (e.g. the SCQM website) about the advances of deep learning in disease prediction gained in this study.

## Notes

### Competing Interest Statement

The authors have declared no competing interest.

### Funding Statement

No funding was obtained for this study.

### Author Declarations

Written informed consent was obtained from all patients and the study was approved by the the regional Ethical committee in Lausanne, Switzerland (ID number 2020-00033).

